# A Social Network Model of the COVID-19 Pandemic

**DOI:** 10.1101/2020.03.23.20041798

**Authors:** Pei Jun Zhao

## Abstract

In the COVID-19 coronavirus pandemic, currently vaccines and specific anti-viral treatment are not yet available. Thus, preventing viral transmission by case isolation, quarantine, and social distancing is essential to slowing its spread. Here we model social networks using weighted graphs, where vertices represent individuals and edges represent contact. As public health measures are implemented, connectivity in the graph decreases, resulting in lower effective reproductive numbers, and reduced viral transmission. For COVID-19, model parameters were derived from the coronavirus epidemic in China, validated by epidemic data in Italy, then applied to the United States. We calculate that, in the U.S., the public is able to contain viral transmission by limiting the average number of contacts per person to less than 7 unique individuals over each 5 day period. This increases the average social distance between individuals to 10 degrees of separation.

## Introduction

From December 31, 2019 to March 18, 2020, the COVID-19 (SARS-CoV2) coronavirus pandemic has infected a reported 209,839 number of people, resulting in 8,778 deaths, in 168 countries around the world, and continue to spread exponentially^1^. While the world awaits the development of vaccines and anti-viral medications, traditional public health measures such as isolation of confirmed or suspected cases, quarantine of contacts, and social distancing of the public are effective measures to reduce viral transmission^2, 3^, that have been implemented in many regions and countries^4, 5^.

COVID-19 is a respiratory infection transmitted by aerosol, droplet, contact, and oral-fecal routes, through networks of social contact. A social network can be visualized as a weighted graph, where vertices represent individuals and edges represent the amount of contact^6^. In the U.S., for example, a pair of individuals can be connected by an average of 6 acquaintances, commonly known as 6 degrees of separation. This small world phenomenon directly affects the spread of COVID-19.

Conceptually, an epidemic is dependent on network connectivity and time^7^. As edges of contact are removed, the average degree of each vertex declines, and connectivity falls. Practically, viral transmission is attenuated by isolation, quarantine and social distancing^8, 9, 10^. These public health measures decrease the number of close contacts, resulting in higher degrees of separation within the public, thereby impeding transmission. This paper examines the level of public health measures needed to contain the COVID-19 pandemic.

### Model

In general, let graph ***g*** represent a weighted network of social contacts. Suppose that it contains *n* vertices, **V**_**1**_ to **V**_**n**_, representing *n* individuals. For a pair of connected vertices **V**_**i**_ and **V**_**j**_, let ***e***_***ij***_ be their common edge with weight *w*_*ij*_. If **V**_**i**_ has degree *k*, then it carries a weighted value of *v*_*i*_=∑ *w*_*ij*_, summed over its *k* edges. Let 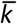 be the average of degree of each vertex and 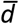 be the average degree of separation of **g**. Then, 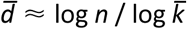.

We begin by modelling viral transmission on ***g***. Suppose individual *i* is infected and had contact with *k* other individuals. Let *w*_*ij*_ be the probability of disease transmission between individuals *i* and *j*. Then *v*_*i*_ is the reproductive number for individual *i*, that is, the average number of secondary infections stemming from *i*. For instance, super-spreading occurs at densely connected vertices, while viral transmission is likely to terminate at sparsely connected vertices.

In a dynamic network, where edges are added and removed, these parameters vary with time corresponding to changes in social behavior. For example, case isolation decreases the degree of **V**_**i**_, while quarantine decreases the degree of vertices connected to **V**_**i**_. Universal precautions such as wearing masks reduces the probability of transmission, *w*_*ij*_.

### Baseline Assumptions

The following assumptions are based on literature review and expert opinion and represent the current knowledge of COVID-19 epidemiology. The values of these parameters often differ by study.

The average generation interval of COVID-19 is about 5 days^11, 12, 13^. (Note that estimates of reproductive numbers are proportional to the assumed generation time.) On average, it take patients 2 days after the start of the infectious period, which can precede symptoms, to seek medical care or report illness^14^. Among sick contacts, 80% are traced by public health and quarantined at home. When individuals practice social distancing such as staying at home, contacts will spend more time with each other by a factor of 1.5. For example, a person who usually spends 12 hours a day with family now spends 18 hours with them. For cases isolated at home, routine precautions reduce household transmission by 50%.

### Parameter Derivation

Using COVID-19 epidemic data from China, the initial epicenter of the outbreak, we will derive parameters for the social network model. First, based on a population study of social mixing patterns in China^15^, the mean number of contacts with unique individuals, over the generation time of COVID-19, is 36, population degree of separation 5.88. Then strict social distancing limits contacts to the household setting and shopping for essential supplies, i.e., 6.3 unique individuals (3.3 in household and 3 in non-household settings), degree of separation 11.5. (See supplementary materials.) Confirmed COVID-19 patients were hospitalized or admitted to isolation centers. Then using open-access daily case reports^16^, we calculate the daily mean reproductive number of the COVID-19 epidemic (Figure 1).

**Figure 1:**
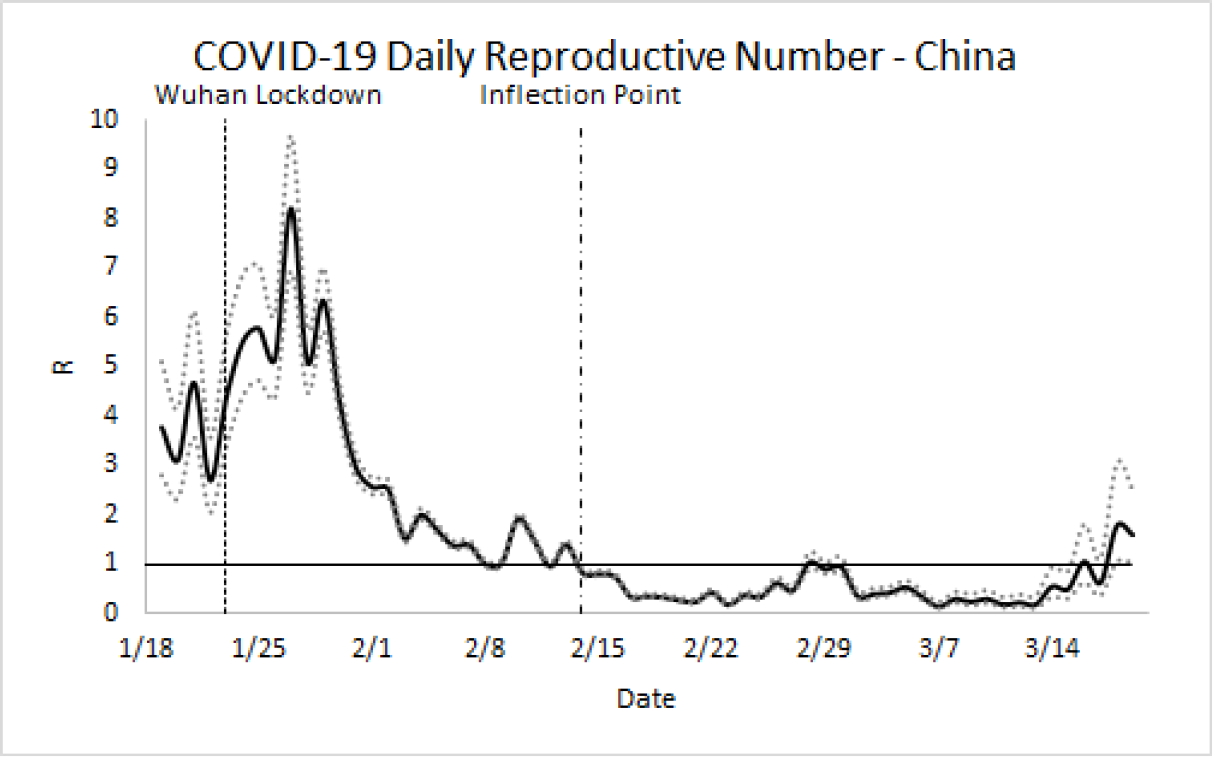
Daily mean reproductive number (R) of COVID-19 in China with 95% confidence intervals. Prior to the lockdown of Wuhan^17^ on January 23, the mean basic reproductive number R_0_ is 3.86. In the few days prior to the lockdown, 5 million people, about a third of the city’s population, left Wuhan for Lunar New Year holidays, and R increased over the following 5 days. With strict social distancing, the epidemic reached an inflection point on February 14 as daily new cases began to consistently decline. An epidemic is contained when R < 1. Since March 15, most new cases are related to travelers returning from abroad.

Prior to intervention, the mean basic reproductive number, R_0_, is estimated to be 3.86. Therefore, the average probability of infection between two contacts is 10.7%, 35.1% for contacts in the household setting versus 8.26% in non-household settings. (See supplementary materials for calculations.) After public health measures are implemented under baseline assumptions, the daily mean reproductive number decreases from 3.86 to 1.99 by social distancing, to 0.79 by isolating all cases at medical facilities, and to 0.41 by quarantine of case contacts, corresponding to the trajectory in Figure 1. During the social distancing period, 81% of new cases are predicted to be through household transmission.

### Model Validation

In Europe, Italy has become the epicenter of the COVID-19 epidemic^18^. From population studies of social patterns in Italy^19^, during the viral generation period, an infected individual is estimated to have 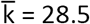 distinct close contacts, 3.6 contacts in the household setting and 24.9 in non-household settings. degree of separation 5.35. The predicted R_0_ is 3.31. (See supplementary materials for calculations.) Again, we calculate the daily mean reproductive numbers based on open-access daily case reports^1^ (Figure 2). The observed R_0_ is 2.99.

**Figure 2:**
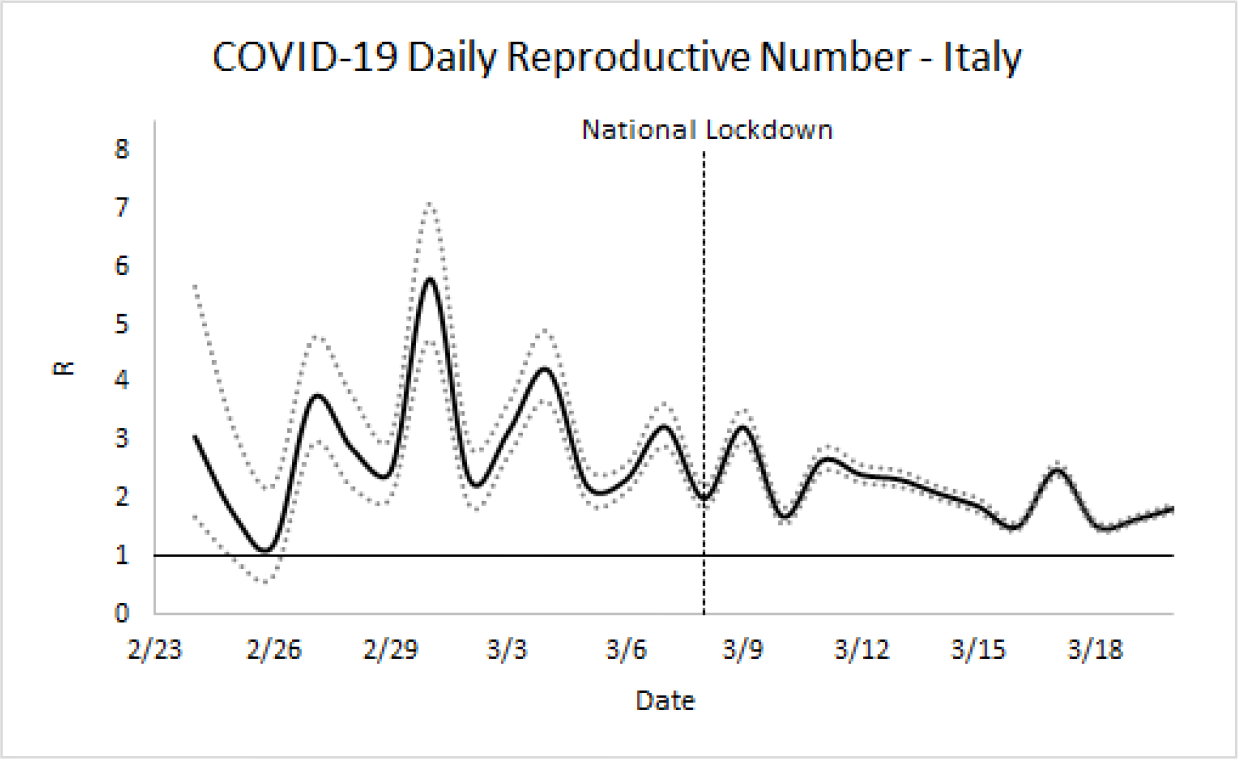
Daily mean reproductive number of COVID-19 in Italy. The estimated mean basic reproductive number R_0_ is 2.99. On March 8, Northern Italy became quarantined, affecting 16 million people^20^. The following day, a national lockdown advisory was issued^21^. On March 11, measures were tightened with closure of most retail services, including fines for violating quarantine orders. On March 21, Italy shut down all non-essential businesses^22^.

**Figure 3:**
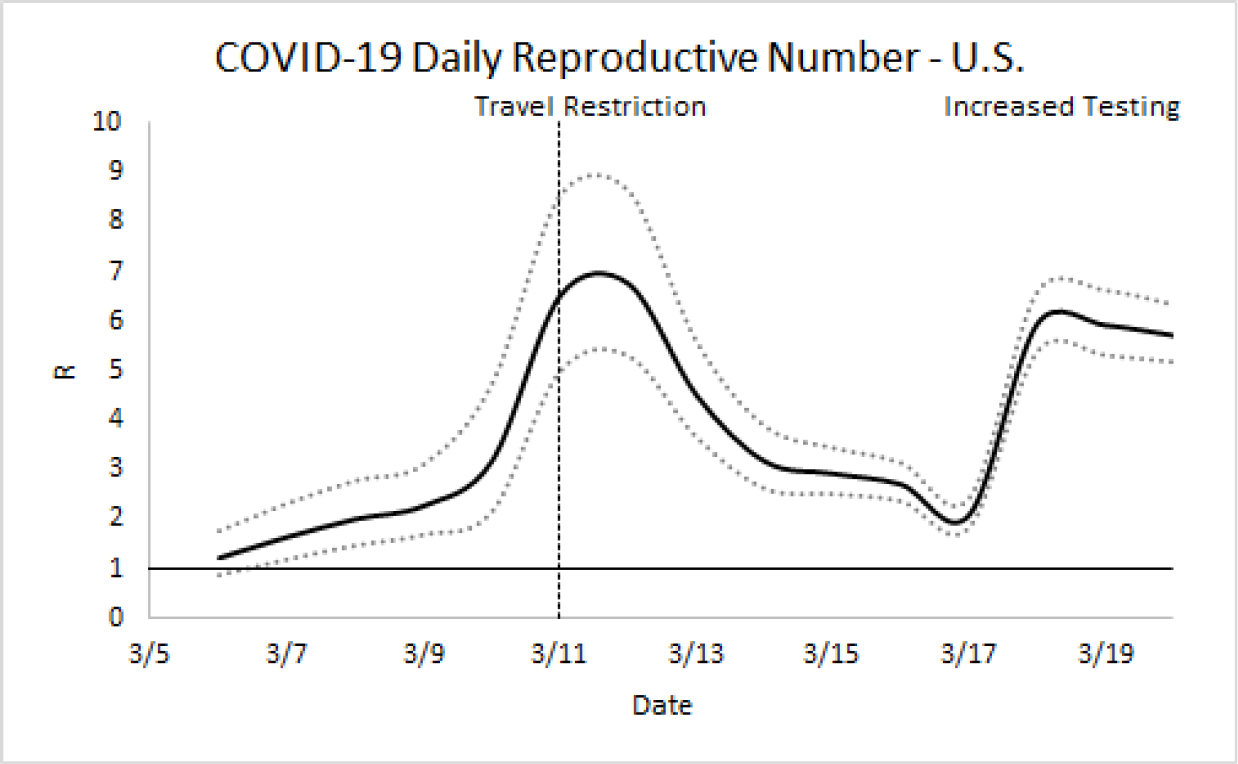
Daily reproductive number of COVID-19 in the United States. The predicted R_0_ for community transmission is about 2.39, that can be seen in the days after the travel restriction to Europe. Since March 17, increased community testing has identified a surge of new cases.

After social distancing, regional, then national quarantine, the daily mean reproductive number has been decreasing. Initial social distancing is estimated to reduce contacts from 28.5 to 11.5, that is, an average of 3.6 contacts in the household setting and 7.9 in the non-household setting, degree of separation 7.35. Furthermore, the entire population is quarantined at home, with moderate to severe cases (20%) hospitalized. Therefore, assuming full compliance with social distancing, the reproductive number is predicted to be 1.03. (See supplementary materials for calculations.) After shutting down all non-essential businesses on March 21, the average number of unique contacts per individual over each viral generation period is expected to be 6.6, leading to a predicted reproductive number of 0.97, to be seen in the coming week.

### Model Prediction

In the U.S., we did not find recent publications on population-level studies of social patterns. Therefore, we assumed that U.S. social patterns are similar to that of Europe^19^. During the viral generation interval, an infected individual is estimated to have 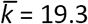 distinct contacts, 2.95 in the household setting and 16.35 in non-household settings, and have 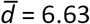 degrees of separation. Thus, the predicted R_0_ = 2.39. (See supplementary materials for calculations.) The initial observed reproductive number became higher than predicted due to travel-acquired infection on top of community transmission. On March 11, the U.S. issued a travel restriction to Europe^5^, that would later include the U.K. After March 17, wide-spread viral testing in the community identified a wave of new cases.

Assume that currently only 70% of cases are diagnosed due to a shortage of test kits^23^. Of confirmed cases, patients with moderate to severe illness (20%) are hospitalized, while patients with mild illness (80%) are isolated at home. Then to contain the epidemic, in addition to public health measures such as contact tracing and quarantine, the average number of unique close contacts per person should be less than 7 over the 5 day generation interval of COVID-19, (3 household contacts and 4 non-household contacts). (See supplementary materials for calculations.) This increases the degrees of separation to 10.

Lastly, we calculate the threshold above which prolonged social distancing measures are needed. Without social distancing, if a single individual is infected with COVID-19, the average probability that any given person will be infected is have 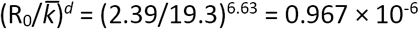, or 1 in 1.03 million. Thus, if 1.03 million people become infected in the U.S., then it is likely that the epidemic will reach every person through networks of social contact, unless social distancing is kept in place until vaccines and anti-viral medications become available.

### Limitations

This paper provides heuristic estimates of COVID-19 epidemiology without knowing the detailed structure of the social network. For large networks, estimates using parameter averages can be robust. There are also numerous factors influencing viral transmission. This simplified model only includes main variables affecting the epidemiology of communicable diseases. Other factors to consider in the future include patient age, sex, pre-existing medical conditions, additional settings of contact such as various workplaces, restaurants, nursing homes, just to name a few. As more factors are taken into consideration, the social network model can be refined to provide more accurate predictions.

## Discussion

The social network model provides a conceptual framework to analyze the COVID-19 pandemic on the level of individuals, that can be used to predict the influence of collective social behavior on overall pandemic trajectories. It shows the importance of social distancing, and the message that to contain the epidemic, every member of the public plays a crucial part in breaking the chain of transmission.

Given the current understanding of COVID-19 viral epidemiology, the epidemic in the U.S. can be controlled by limiting the average number of contacts per person to 7 unique individuals over each 5 day period. For an average American family, the 7 contacts might be a spouse, two children, a friend, a neighbor, a colleague, and a cashier during grocery shopping. The time from the beginning of collective social action to the inflection point of the epidemic is about 3 viral generation intervals, totaling 2-3 weeks. Once daily new cases are decreasing, social distancing measures can be gradually relaxed, in phases, to prevent rebound of viral transmission.

After social distancing becomes widely implemented and most people stay at home, the majority of new cases is expected to be from household transmission. One way to further reduce the epidemic is to bring all patients with confirmed COVID-19 to medical facilities to prevent the spread of infection at home, as was the practice in China, where moderate to severe cases were hospitalized while mild cases were kept in isolation centers until they were no longer infectious on viral nucleic acid testing. This is also the standard of care for other communicable diseases such as tuberculosis, where clinically stable patients are discharged home after three negative induced sputum samples.

Finally, the social network model of COVID-19 transmission is expandable and extendable. This paper provides a simplified version using population averages for parameters. Researchers who have access to patient-level data can substitute these average values by their respective probability distributions to simulate stochastic viral transmission.

## Data Availability

Data used in this paper are open access and can be found in the cited references.

